# Age and Sex Differences of Virtual Reality Pain Alleviation Therapeutic During Pediatric Burn Care: A Randomized Clinical Trial

**DOI:** 10.1101/2024.01.26.24301834

**Authors:** Katarina Jones, Megan Armstrong, John Luna, Rajan K Thakkar, Renata Fabia, Jonathan I Groner, Dana Noffsinger, Ai Ni, Bronwyn Griffin, Henry Xiang

**Affiliations:** College of Medicine, Northeast Ohio Medical University, 4209 St, OH-44, Rootstown, OH 44272, USA; Center for Pediatric Trauma Research, The Abigail Wexner Research Institute, Nationwide Children’s Hospital, 700 Children’s Drive, Columbus, OH 43205, USA; Center for Injury Research & Policy, The Abigail Wexner Research Institute, Nationwide Children’s Hospital, 700 Children’s Drive, Columbus, OH 43205, USA; IT Research and Innovation, The Abigail Wexner Research Institute, Nationwide Children’s Hospital, 700 Children’s Drive, Columbus, OH 43205, USA; Trauma and Burn Program, Nationwide Children’s Hospital, 700 Children’s Drive, Columbus, OH 43205, USA; College of Medicine, The Ohio State University, 370 West 9th Avenue, Columbus, OH 43210, USA; Division of Biostatistics, The Ohio State University College of Public Health, 1841 Neil Avenue, Columbus, OH 43210, USA; NHMRC Centre of Research Excellence-Wiser Wound Care, Menzies Health Institute of Queensland, Griffith University Brisbane, N48 Nathan Campus, Brisbane, QLD, Australia

**Keywords:** Virtual Reality, Burn, Pediatric, Pain

## Abstract

Virtual reality (VR) effectively alleviates pain for pediatric patients during many medical care procedures, such as venipuncture and burn wound care. Whether VR pain alleviation therapeutics (VR-PAT) differ by a patient’s age or sex remains unresolved. This randomized clinical trial evaluated how age and sex affect VR pain alleviation during dressing care for pediatric burns. Ninety patients aged 6-17 years (inclusive) with burn injuries were recruited from an outpatient burn clinic of an American Burn Association-verified pediatric burn center. Before randomization, expectations of VR helpfulness and need were assessed on a visual analog scale (VAS, 0-100). Participants were randomly assigned to active or passive VR for one burn dressing change. Immediately following the dressing change, participants self-reported pain and the time spent thinking about pain and rated the VR features on the degrees of realism experienced, pleasure/fun, and perceived engagement level. Path analyses assessed how these VR features were interrelated and how they affected self-reported pain by age and sex. Patients aged 6–9 years reported higher mean expectations of VR helpfulness and need (mean=73.6 and 94.5, respectively) than 10–12-year-olds (mean=55.7 and 84.2, respectively) and 13-17-year-olds (mean=68.6 and 77.4, respectively). The path analysis indicated VR engagement and fun were significantly correlated (p-value <0.05). VR engagement significantly negatively impacted overall pain scores during burn dressing (coefficient=-0.45, −0.41; p-value <0.05) and significantly positively impacted time thinking of pain (coefficient=0.38, 0.32; p-value <0.05). Younger patients had the highest expectations of VR pain alleviation’s helpfulness and need. VR game realism, fun, and engagement features were not statistically different between age groups and sexes. VR engagement and thinking of pain during burn dressing significantly positively affected self-reported pain (p-value <0.05), suggesting an analgesic mechanism beyond distraction alone. Younger patients benefited more from VR pain alleviation therapeutics than older patients.

## Introduction

In recent years, several nonpharmacological methods for pain alleviation have been explored, including VR (virtual reality). VR has been established as an effective method to distract pediatric patients and alleviate pain during particularly painful medical procedures.^1^ Although VR has been associated with some short-term side effects due to simulator sickness (including nausea, eye strain, dizziness, and headache),^2^ VR is generally considered a promising nonpharmacological approach for further research and clinical use.^3^ In particular, VR pain alleviation therapeutics are very effective among pediatric patients.^4^ For example, VR reduces pain in younger children during burn hydrotherapy sessions.^5^ While VR has been used for many clinical pain conditions^6–10^ that include wide age ranges and both sexes, whether VR pain alleviation differs according to a patient’s age or sex remains unresolved. Whether and how a patient’s age or sex may influence VR pain alleviation therapeutics during medical procedures needs further investigation.

Burn injury is among the top ten leading causes of death and unintentional injury in children.^11^ Over 67,000 US children sustained nonfatal burn injuries in 2020.^12^ Patients with severe burns experience background pain that exists during rest and procedural pain from wound care procedures adds to this existing pain.^13^ Painful burn dressing experiences can stress patients significantly enough to affect post-injury health outcomes.^14^ Pharmacological methods, such as opioids, are often used as the standard treatment to address this pain;^13^ however, opioid tolerance and dependence may increase over time.^15^ Due to the current US opioid abuse epidemic, nonpharmacological approaches to alleviate pain are particularly needed.

Pain is a subjective experience, with pain perception and rating differing based on sex.^16,17^ However, many studies use experimental pain conditions, ^18–20^ which are different from pain caused by medical conditions such as burn injury. Around puberty, sex differences in pain emerge, with adolescent girls reporting more pain than adolescent boys.^21^ In addition, pediatric patients tend to process pain in more complex ways as they age.^22^

This study aimed to (1) assess whether age or sex impacted how our VR pain alleviation therapeutics (VR-PAT) affected pain during pediatric burn dressings, (2) evaluate whether different perceptions of VR-PAT (game realism, fun, engagement) correlated with pain alleviation, and (3) evaluate whether prior expectations of VR efficacy significantly influenced pain alleviation during burn dressing changes. Our central hypothesis was that VR features (game realism, fun, engagement) could be rated differently depending on the age or sex of the patients. In turn, these key VR features could significantly impact the effectiveness of VR pain alleviation therapeutics.

## Methods

### Data Source

Data used in these analyses were collected from a randomized controlled trial designed to assess VR-PAT efficacy to manage pain during pediatric burn dressing changes in an outpatient clinic. The study design and protocols are described in a prior publication.^23^ Patients were randomly assigned to an active VR group (immersive VR with gameplay), passive VR group (same immersive environment without gameplay), or standard care control group (using conventional distraction methods available in the clinic, such as iPads, music, books, and/or talking). Self-reported, observed, and burn wound data were collected during one scheduled dressing change within a six-day median since the initial injury at the outpatient burn clinic of an American Burn Association (ABA)-verified pediatric burn center.

The Institutional Review Board of the children’s hospital reviewed and approved the study. Written informed consent from one legal guardian and written assent from children nine years and older were obtained before beginning study measures. This study was registered at ClinicalTrials.gov, with the identifier NCT04544631.

### Study Population

Inclusion criteria were children aged 6 to 17 years (inclusive) treated at the outpatient burn clinic of an ABA-verified pediatric burn center between December 2016 and January 2019 and spoke English as their primary language. Exclusion criteria were (1) a severe burn on the face or head that prevented VR use; (2) cognitive or motor impairment that prevented study measure administration; (3) visual or hearing impairments that prevented VR interaction; or (4) did not have a legal guardian present to give consent. These inclusion and exclusion criteria (including age range) were designed to be most appropriate for VR immersion and administration of study measures.

### Study Procedures

After informed consent but before randomization, a trained researcher conducted a pre-intervention survey, asking patients about their expectations of VR distraction effectiveness. These questions were “How much would you like to have something fun to do during the dressing change?” and “How much do you think it would help with your pain during the dressing change?” These questions were assessed using a 100 mm VAS, where a higher number indicated more expectation of effectiveness. Guardians were asked whether the child took any pain medications within six hours before the burn wound care appointment at the outpatient clinic.

Following the pre-intervention survey, participants were randomized to active VR, passive VR, or the control group. The randomization scheme used a 1:1:1 allocation ratio and was balanced by sex and stored in a Research Electronic Data Capture (REDCap) database.^24,25^

Immediately following the burn wound care, another trained researcher blinded to the patient’s intervention group conducted a post-intervention survey. Patients were asked to report their overall pain, worst pain, and time spent thinking about pain during the burn dressing change procedures using the VAS (0-100, with higher scores indicating more pain or more time thinking about pain). Participants in the VR groups were asked to rate the VR game’s realism (“How realistic did you feel about it?”), fun (“How much fun did you have with the VR?”), engagement (“How engaging did you think it was?”) using the 0-100 VAS, with higher scores indicating more realism, fun, or engagement.

Patient demographic information and burn injury characteristics were obtained from each patient’s electronic medical record. For this analysis, demographic variables included sex (male, female) and date of birth (used to calculate age and grouped as 6-9, 10-12, and 13-17 years). Burn injury characteristics included the percentage of total body surface area (TBSA) (<1%, 1.0-4.9%, 5.0-25.0%) and burn degree (first, second, third). Nurses involved in burn wound care were asked to report the degree of healing of the wound using a Likert scale (minimal healing present, partially healed burn [<50% healed], mostly healed burn [>50% healed], or completely healed).

### Outcome Measures

The primary outcome was a self-reported overall pain score during the burn dressing procedure. After the burn wound care, a trained researcher asked the patients about their overall pain during the whole burn dressing change using a VAS (range 0-100, with higher scores indicating more pain).

### Statistical Analysis

Based on published randomized controlled trial studies,^23,26^ we assumed a medium effect size (f^2^ = 0.15) of the active VR. Using two tails and α = 0.05, we estimated a fixed-effect linear regression model offering power >0.80 with a total sample size of 90 children. We aimed to recruit 30 participants for each group (active VR, passive VR, and standard-of-care control group) to ensure adequate study power.

As this study aimed to determine how overall pain score differed by age and sex, and we postulated features of the core VR experience (VR game realism, fun, engagement) would vary by age and could significantly impact the effectiveness of VR pain alleviation therapeutics, the standard-of-care group was excluded from the final statistical analysis. We first compared the frequency of active vs. passive VR assignment, burn injury characteristics (%TBSA, burn degree, healing degree), and any pain medication used within six hours before the burn wound care by study participants’ sex and age categories (6-9, 10-12, 13-17 years) to ensure comparability of patients in active vs. passive VR groups. We then compared the numerical overall pain score, VR expectation, and core VR experience (VR game realism, fun, engagement) using mean, median, lower quartile, and upper quartile by sex and age categories. P-values of the statistical difference test were calculated from nonparametric Wilcoxon rank sum tests.

We conducted path analysis modeling to estimate the hypothesized relationships among key VR features (VR game realism, fun, engagement) and their impact on the primary outcome (patient self-reported pain score) via the intermediate variable “time thinking about pain” during burn dressing. An extension of multiple regression, path analysis estimates the magnitude and significance of hypothesized connections between variable sets.^27^ Path analysis also allows simultaneous assessment of multiple independent variables’ direct and indirect effects on the primary outcome variable (self-reported overall pain score during burn dressing). A direct effect is observed when an independent variable directly influences a dependent variable. In contrast, an indirect effect occurs when an independent variable affects a dependent variable through a mediating variable. Standardized path coefficients with corresponding p-values could help determine independent variables’ significant direct and indirect effects on the study’s primary outcome variable.

We used SAS version 9.4 to conduct data analyses.^28^ Statistical significance was set at α < 0.05, and all tests were two-tailed.

## Results

Four hundred twelve children were screened for eligibility, with 240 eligible and 95 recruited into the study (**Fig. 1**). Of those recruited, 90 completed the pre-intervention survey and were randomized. For this analysis, the 29 patients randomized to the control group were excluded to assess age and sex differences in their VR features rating, self-reported time thinking about pain, and overall pain intensity score during burn dressing care, leaving 61 subjects for the current analysis.

**Fig 1:**
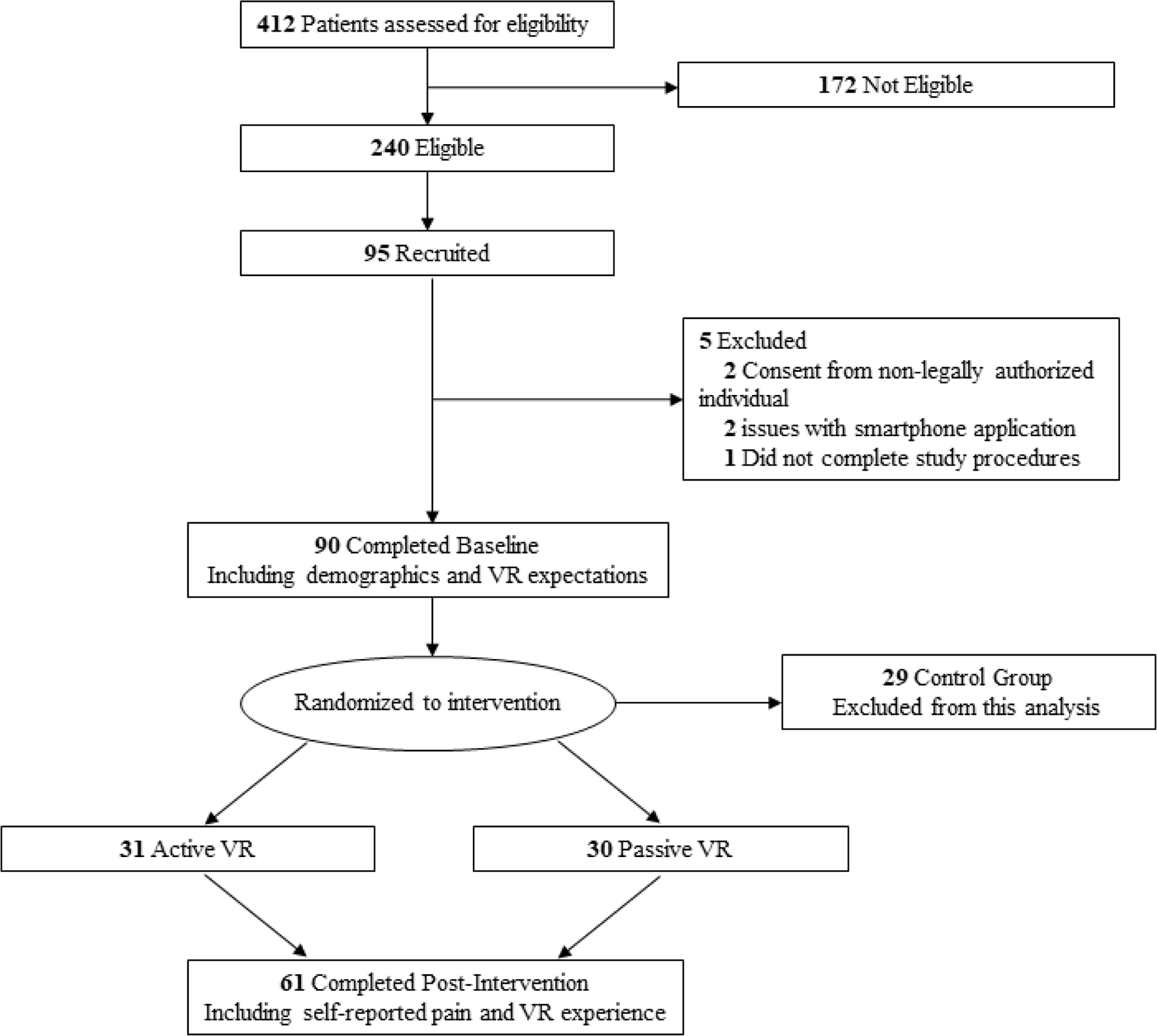
CONSORT Flow Diagram of patient screening, recruitment, and study procedures.

Burn characteristics and pain medication used within six hours before the burn dressing care by age and sex were reported in **Table 1**. Age and sex did not significantly differ by VR group assignment, TBSA burned, burn degree, healing degree, or pain medication within six hours of dressing (p-value <0.05).

**Table 1.** Demographics and burn characteristics of participants aged 6-17 years who used VR-PAT during burn dressing changes (N=61)

|  | Sex |  | *P* | Age category |  |  | *P* |
| --- | --- | --- | --- | --- | --- | --- | --- |
|  | Male<br>(n,%) | Female<br>(n,%) |  | 6-9 yrs<br>(n,%) | 10-12 yrs<br>(n,%) | 13-17 yrs<br>(n,%) |  |
| **VR group** |  |  | 0.89 |  |  |  | 0.90 |
| Active VR | 16 (50.0%) | 15 (51.7%) |  | 10 (32.3%) | 9 (29.0%) | 12 (38.7%) |  |
| Passive VR | 16 (50.0%) | 14 (48.3%) |  | 11 (36.7%) | 9 (30.0%) | 10 (33.3%) |  |
| **TBSA burned** |  |  | 0.31 |  |  |  | 0.57 |
| Missing | 0 (0.0%) | 1 (3.5%) |  | 0 (0.0%) | 1 (5.6%) | 0 (0.0%) |  |
| <1% | 12 (37.5%) | 11 (37.9%) |  | 6 (28.6%) | 6 (33.3%) | 11 (50%) |  |
| 1.0-4.9 % | 15 (46.9%) | 16 (55.2%) |  | 13 (61.9%) | 9 (50.0%) | 9 (40.9%) |  |
| 5-25% | 5 (15.6%) | 1 (3.5%) |  | 2 (9.5%) | 2 (11.11%) | 2 (9.1%) |  |
| **Burn degree** |  |  | 0.09 |  |  |  | 0.20 |
| Missing | 0 (0.0%) | 1 (3.5%) |  | 0 (0.0%) | 1 (5.6%) | 0 (0.0%) |  |
| Second degree | 28 (87.5%) | 28 (87.5%) |  | 18 (85.7%) | 16 (88.9%) | 22 (100%) |  |
| Third degree | 4 (12.5%) | 0 (0.0%) |  | 3 (14.3%) | 1 (5.6%) | 0 (0.0%) |  |
| **Heal degree** |  |  | 0.95 |  |  |  | 0.52 |
| Minimal | 13 (40.6%) | 10 (34.5%) |  | 10 (47.6%) | 2 (33.3%) | 7 (31.8%) |  |
| Partially healed (<50%) | 9 (28.1) | 8 (27.6%) |  | 6 (28.6%) | 4 (22.2%) | 7 (31.8%) |  |
| Mostly healed (>50%) | 7 (21.9%) | 8 (27.6%) |  | 2 (9.5%) | 6 (33.3%) | 7 (31.8%) |  |
| Completely healed | 3 (9.4%) | 3 (10.3%) |  | 3 (14.3%) | 2 (11.1%) | 1 (4.6%) |  |
| **Pain medication within 6 hours before dressing change** |  |  | 0.30 |  |  |  | 0.11 |
| Missing | 0 (0.0%) | 2 (6.9%) |  | 0 (0.0%) | 2 (11.1%) | 0 (0.0%) |  |
| No | 20 (62.5%) | 18 (62.1%) |  | 12 (57.1%) | 13 (72.2%) | 13 (59.1%) |  |
| Yes | 12 (37.5%) | 9 (31.0%) |  | 9 (42.9%) | 3 (16.7%) | 9 (40.9%) |  |

TBSA, total body surface area; VR PAT, virtual reality pain alleviation therapeutics; yrs, years

The self-reported experience of 61 patients utilizing VR pain alleviation therapeutics is reported in **Table 2**. Expectations significantly differed by age and patients who expected VR to be helpful and necessary for pain alleviation (p-value = 0.05, 0.02, respectively). Younger patients 6–9 years had higher expectations of VR helpfulness and pain alleviation needs (mean = 73.6 and 94.5, respectively) than those 10–12 years (mean = 55.7 and 84.2, respectively) and 13-17 years (mean = 68.6 and 77.4, respectively). However, VR game realism, fun, or engagement did not significantly vary by age or sex. In addition, VR expectations or overall pain scores did not significantly differ by sex. However, overall pain scores differed by age, with the youngest patients reporting the highest pain (mean = 40.4), compared to those 10-12 (mean = 35.9) and 13-17 (mean = 16.0; p-value=0.04).

**Table 2.**
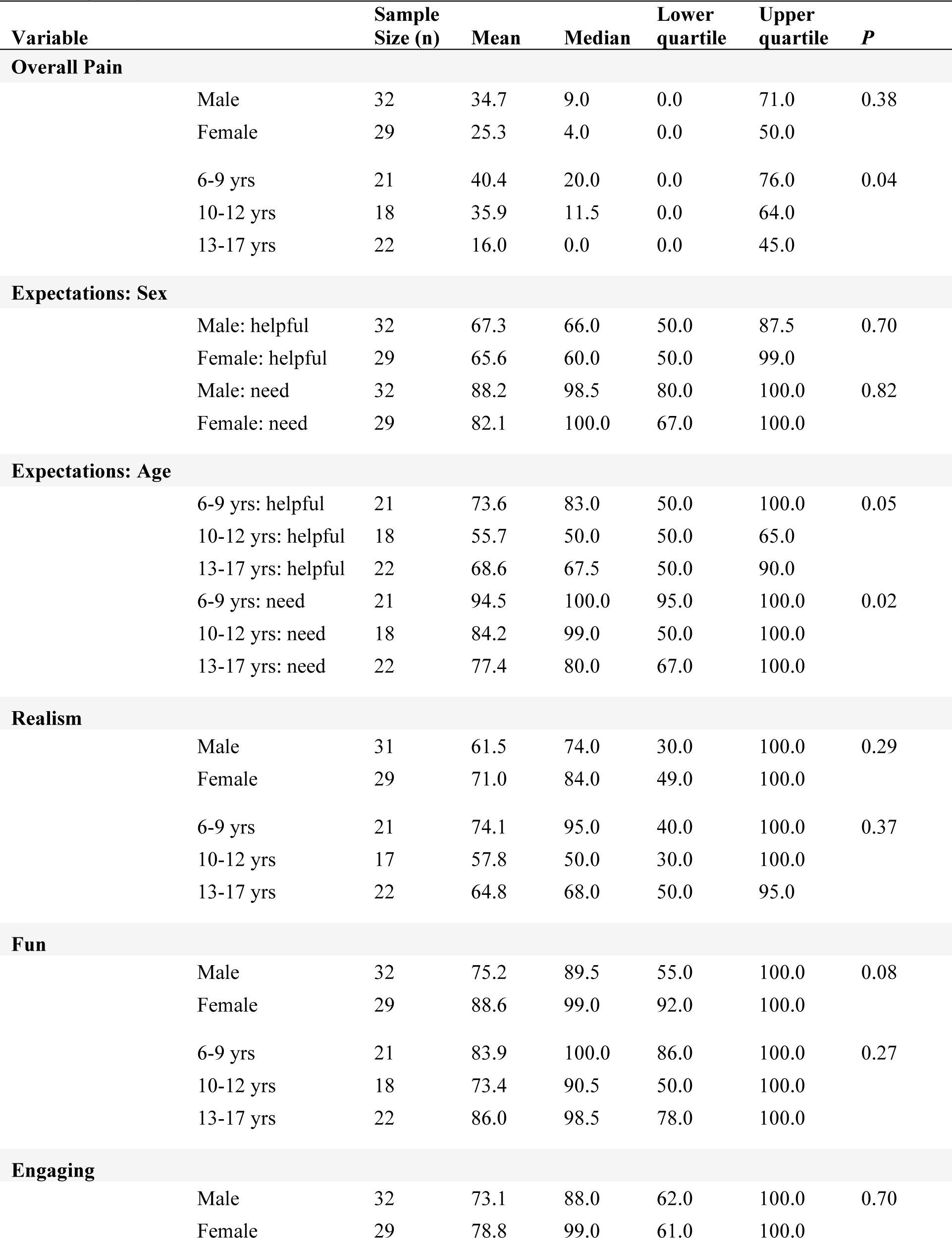
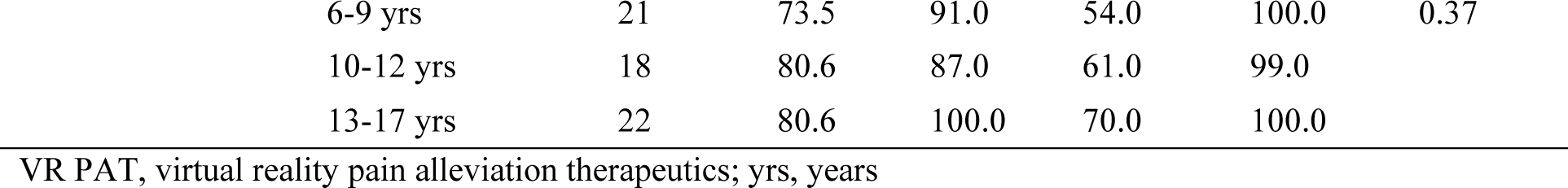
Self-reported experience of participants aged 6-17 years who used VR-PAT during burn dressing changes (N=61)

| Variable |  | Sample Size (n) | Mean | Median | Lower quartile | Upper quartile | P |
| --- | --- | --- | --- | --- | --- | --- | --- |
| Overall Pain |  |  |  |  |  |  |  |
|  | Male | 32 | 34.7 | 9.0 | 0.0 | 71.0 | 0.38 |
|  | Female | 29 | 25.3 | 4.0 | 0.0 | 50.0 |  |
|  | 6-9 yrs | 21 | 40.4 | 20.0 | 0.0 | 76.0 | 0.04 |
|  | 10-12 yrs | 18 | 35.9 | 11.5 | 0.0 | 64.0 |  |
|  | 13-17 yrs | 22 | 16.0 | 0.0 | 0.0 | 45.0 |  |
| Expectations: Sex |  |  |  |  |  |  |  |
|  | Male: helpful | 32 | 67.3 | 66.0 | 50.0 | 87.5 | 0.70 |
|  | Female: helpful | 29 | 65.6 | 60.0 | 50.0 | 99.0 |  |
|  | Male: need | 32 | 88.2 | 98.5 | 80.0 | 100.0 | 0.82 |
|  | Female: need | 29 | 82.1 | 100.0 | 67.0 | 100.0 |  |
| Expectations: Age |  |  |  |  |  |  |  |
|  | 6-9 yrs: helpful | 21 | 73.6 | 83.0 | 50.0 | 100.0 | 0.05 |
|  | 10-12 yrs: helpful | 18 | 55.7 | 50.0 | 50.0 | 65.0 |  |
|  | 13-17 yrs: helpful | 22 | 68.6 | 67.5 | 50.0 | 90.0 |  |
|  | 6-9 yrs: need | 21 | 94.5 | 100.0 | 95.0 | 100.0 | 0.02 |
|  | 10-12 yrs: need | 18 | 84.2 | 99.0 | 50.0 | 100.0 |  |
|  | 13-17 yrs: need | 22 | 77.4 | 80.0 | 67.0 | 100.0 |  |
| Realism |  |  |  |  |  |  |  |
|  | Male | 31 | 61.5 | 74.0 | 30.0 | 100.0 | 0.29 |
|  | Female | 29 | 71.0 | 84.0 | 49.0 | 100.0 |  |
|  | 6-9 yrs | 21 | 74.1 | 95.0 | 40.0 | 100.0 | 0.37 |
|  | 10-12 yrs | 17 | 57.8 | 50.0 | 30.0 | 100.0 |  |
|  | 13-17 yrs | 22 | 64.8 | 68.0 | 50.0 | 95.0 |  |
| Fun |  |  |  |  |  |  |  |
|  | Male | 32 | 75.2 | 89.5 | 55.0 | 100.0 | 0.08 |
|  | Female | 29 | 88.6 | 99.0 | 92.0 | 100.0 |  |
|  | 6-9 yrs | 21 | 83.9 | 100.0 | 86.0 | 100.0 | 0.27 |
|  | 10-12 yrs | 18 | 73.4 | 90.5 | 50.0 | 100.0 |  |
|  | 13-17 yrs | 22 | 86.0 | 98.5 | 78.0 | 100.0 |  |
| Engaging |  |  |  |  |  |  |  |
|  | Male | 32 | 73.1 | 88.0 | 62.0 | 100.0 | 0.70 |
|  | Female | 29 | 78.8 | 99.0 | 61.0 | 100.0 |  |

|  |  |  |  |  |  |  |
| --- | --- | --- | --- | --- | --- | --- |
| 6-9 yrs | 21 | 73.5 | 91.0 | 54.0 | 100.0 | 0.37 |
| 10-12 yrs | 18 | 80.6 | 87.0 | 61.0 | 99.0 |  |
| 13-17 yrs | 22 | 80.6 | 100.0 | 70.0 | 100.0 |  |

VR PAT, virtual reality pain alleviation therapeutics; yrs, years

The standardized direct and indirect effects of age and sex on VR pain scores calculated from path analysis models were reported in **Table 3**. VR engagement significantly negatively affected pain scores in both models (coefficient = −0.45, −0.41, p-value <0.05, respectively). Time thinking of pain during burn dressing care significantly affected both models (coefficient = 0.38, 0.32 p-value<0.05, respectively), proving a significant role of distraction in pain alleviation. In the path analysis model by patient age, age significantly negatively impacted the self-reported overall pain score (coefficient = −0.30, p <0.05), suggesting that VR pain alleviation decreased as the patient’s age increased.

**Table 3.** Standardized direct and indirect effects^a^ of sex, age, and virtual reality features, on self-reported overall pain score (0-100) during pediatric burn dressing changes (6-17 years, N=61)

| **Effect** | **Direct** | **Indirect** | **Total** |
| --- | --- | --- | --- |
| Sex (male=1; female=0) | 0.10 | 0.03 | 0.12 |
| VR fun (0-100) | -0.12 | -0.06 | -0.18 |
| VR engaging (0-100) | -0.39<sup>b</sup> | -0.05 | -0.45<sup>b</sup> |
| Time thinking of pain during burn dressing (0-100) | 0.37<sup>b</sup> |  | 0.38<sup>b</sup> |
| Age (6-17 years) | -0.20<sup>b</sup> | -0.10 | -0.30<sup>b</sup> |
| VR fun (0-100) | -0.17 | -0.05 | -0.22<sup>b</sup> |
| VR engaging (0-100) | -0.37<sup>b</sup> | -0.04 | -0.4<sup>b</sup> |
| Time thinking of pain during burn dressing (0-100) | 0.32<sup>b</sup> |  | 0.32<sup>b</sup> |

<sup>a</sup>Two separate path analysis models were conducted for age and sex
<sup>b</sup> p<0.05
VR, virtual reality

The association between multiple variables and patient self-reported pain by sex and age is shown in **Fig 2A-B**. VR engagement and fun were significantly correlated (coefficient = 0.63, p-value <0.05). VR engagement and overall pain score were significantly negatively correlated by sex and age (coefficient = −0.39, −0.37; p-value <0.05). Time spent thinking about pain during burn dressing care had a significant positive correlation with pain score by sex and age (coefficient = 0.38, 0.32; p-value <0.05). The patient’s sex and reporting of having fun were significantly negatively correlated (coefficient =-0.24, p-value<0.06), suggesting that female patients had more fun than male patients.

**Fig 2.**
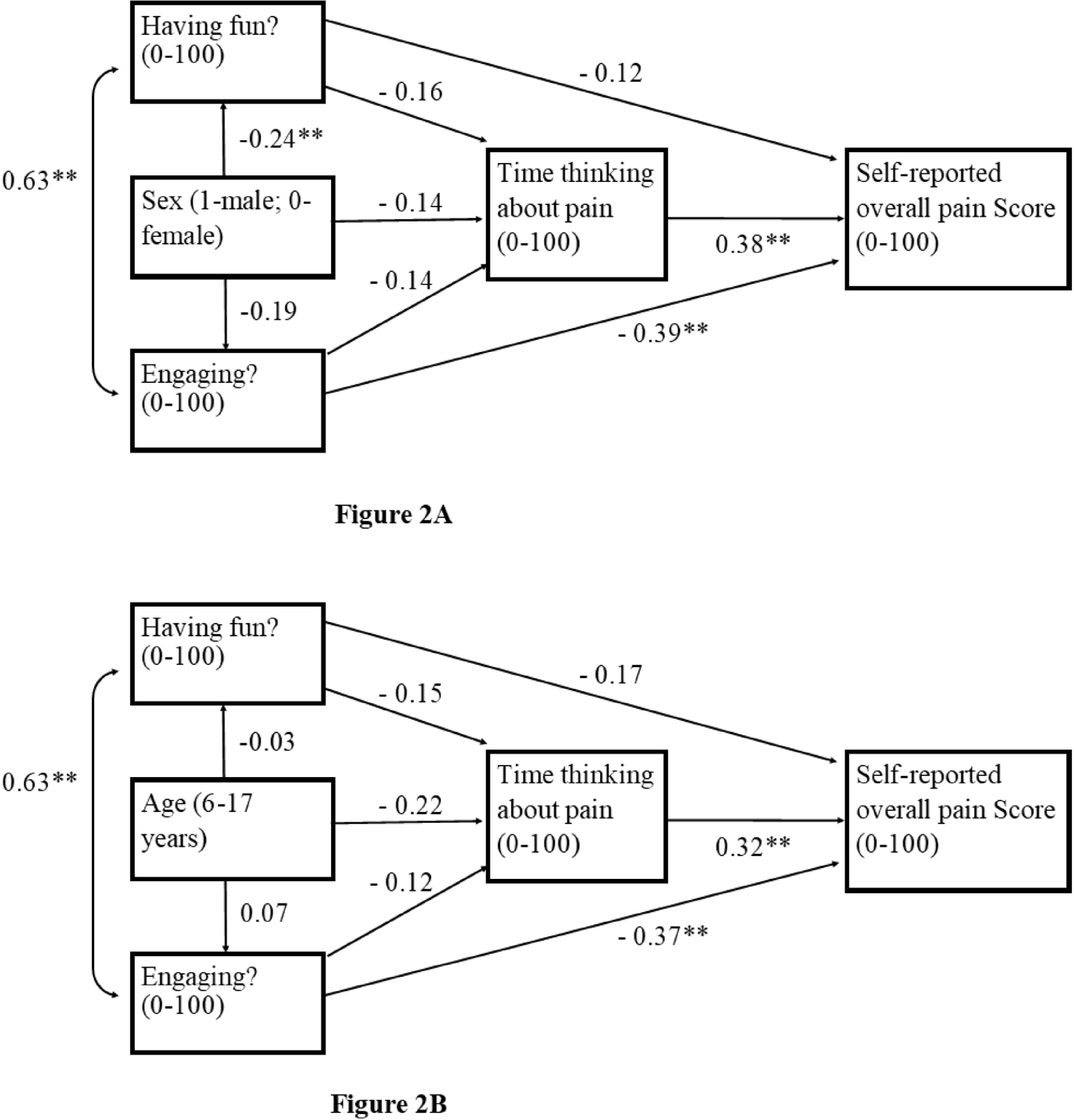
Postulated sex path model (**2A**) and age path model (**2B**) for virtual reality pain effect during dressing changes. Association between multiple variables and patient self-reported pain is shown by arrows extending from each variable to self-reported overall pain score (0-100). Numbers on each arrow are standardized path coefficients. The higher the coefficient indicates a stronger association where ** indicates p <0.05

## Discussion

Our study investigated how patient expectation of VR-PAT, need for VR, and key VR features (VR game realism, fun, engagement) could affect the self-reported pain intensity score during pediatric burn dressing care across age ranges and by sex. VR realism, engagement, and fun did not significantly vary by sex or age. VR engagement correlated with greater fun and lower overall pain scores across age and sex, while time thinking about pain during burn dressing care correlated significantly with increased self-reported overall pain scores. Younger patients expected the VR experience to be more necessary and helpful in reducing pain, and female patients reported more fun with VR than male patients.

Our results suggest nearly all pediatric patients, regardless of age and sex, could benefit from the VR pain alleviation therapeutics and experience a significantly reduced overall pain intensity during burn dressing procedures. However, our findings differ from a previous study where younger children had more “fun” with VR.^29^ Future research could include investigations on how “fun” by age differs in VR pain alleviation therapeutics.

In our study, VR engagement was significantly correlated with fun, and fun and engagement significantly reduced pain intensity scores in the path analysis model assessing patient age, VR fun and engagement, and time thinking about pain during burn dressing care. Compared to a non-interactive VR experience, interactive and engaging VR was considered more fun,^30^ and VR that is demanding and engages more mental load reduced the intensity and unpleasantness of nociceptive stimuli.^31^ In addition, patients who reported more fun also reported lower pain intensity scores,^32^ which could be because children are generally more responsive to VR pain alleviation therapeutics that involve play.^33^ Subjective pain intensity ratings (via functional magnetic brain imaging) significantly decreased during VR immersion, followed by decreased pain-related brain activity and higher inhibition of the pain matrix in the brain.^34,35^ These brain imaging data suggest a neuro-mechanism for the nonpharmacological analgesia achieved through VR pain alleviation. Since 1965, the presiding theory for how VR alleviates pain is that VR competes for a fixed amount of the user’s attention, replacing nociceptive input with pleasant sensory input through descending inhibitory pathways.^36^ Our study provides further evidence that VR engagement, which correlates with fun, could alleviate pain on a neurological level beyond mere distraction (i.e., time thinking about pain during burn dressing care).

Engaged patients reported lower pain scores and patients who spent more time thinking about pain reported higher pain scores. Pain requires attention,^37^ so the attention-grabbing nature of VR could leave less attention available to process incoming pain signals.^38^ Previous trials have indicated that VR reduces the time patients think about pain.^31,39^ By engaging their attention with the VR pain alleviation therapeutics, patients could have spent less time thinking about pain during the burn dressing care and, in return, subjectively experienced less pain.

Younger patients had higher expectations of VR pain alleviation therapeutics’ helpfulness and need. Pain expectations can influence how children perceive, express, and respond to pain.^40^ In a previous study of expectations going into a gaming experience, participants with favorable expectations of the distraction reported more enjoyment and engagement, and lower pain intensity scores.^41^ The youngest group reported the highest pain intensity scores in our study. However, the age path analysis model suggested that age had a significant, negative direct, and overall impact on self-reported pain intensity. Our findings were consistent with a prior study that examined VR distraction during dental anesthesia, in which the youngest patients reported the highest pain scores.^42^ When compared to video games, head-mounted display VR raised the pain tolerance and threshold in children over ten but did not significantly affect participants under ten,^43^ which could be due to younger children’s higher sensitivity to noxious stimuli.^44^ Overall, age differences in VR pain alleviation therapeutics remain largely unresolved.

A strength of our study was that demographics and burn characteristics of our patients were comparable, i.e., the randomized patients did not significantly differ by age, sex, or burns treated. One limitation was that the effect of VR pain alleviation therapeutics was only measured once, instead of during repeated burn dressing care over time. However, previous studies found that VR is effective for multiple treatments.^45,46^ Another limitation of our study is that VR experience and pain intensity scores were subjective and self-reported by the patients, which is a common weakness in the pain research field. Objective measures such as neuro-imaging biomarkers should be developed and tested in future research.

## Conclusions

VR was similarly fun, engaging, and realistic across age and sex but less effective in alleviating pain in older children. VR engagement and time spent thinking about pain during burn dressing had a significant direct positive effect on self-reported pain, suggesting an analgesic mechanism beyond distraction alone. Future development and research of VR pain alleviation therapeutics should include key VR features that could significantly impact VR pain alleviation effectiveness.

## Data Availability

All data produced in the present study are available upon reasonable request to the authors.

## Acknowledgements

We thank Beth Reichard, Scientific Writing Program Coordinator at the Abigail Wexner Research Institute for editing the manuscript. Thanks to nurses and staff of the Burn Center at Nationwide Children’s Hospital for coordination and support of the clinical trial.

## Author Contributions CREDIT Statement

Katarina Jones: Results Interpretation, Writing – Original Draft

Megan Armstrong: Project administration, Results Interpretation, Writing – Review and Editing

John Luna: VR-PAT Development, Results Interpretation, Writing – Review and Editing

Dana Noffsinger: Conceptualization, Implementation, Results Interpretation, Writing – Review and Editing

Ai Ni: Methodology, Data Analysis Guidance, Results Interpretation, Writing – Review and Editing

Bronwyn Griffin: Results Interpretation, Writing – Review and Editing

Rajan Thakkar, Renata Fabia, Jonathan Groner, and Henry Xiang: Conceptualization, Supervision, Resources, Results Interpretation, Writing – Review and Editing, Funding Acquisition

